# SARS-CoV-2 B.1.1.7 lineage rapidly spreads and overwhelms R.1 lineage in Japan: serial and stationary observation in a community

**DOI:** 10.1101/2021.06.30.21259820

**Authors:** Yosuke Hirotsu, Masao Omata

**Author notes:** Corresponding author: Yosuke Hirotsu, Genome Analysis Center, Yamanashi Central Hospital, 1-1-1 Fujimi, Kofu, Yamanashi, Japan.

## Abstract

**Background:** The severe acute respiratory syndrome coronavirus 2 (SARS-CoV-2) circulates in the world and acquires mutations during evolution. To identify the new emergent variants, the surveillance of the variants of concern (VOC) and variants of interest (VOI) is ongoing. This study aimed to determine how the transition of viral lineage occurred by stationary genome analysis in Yamanashi, Japan.

**Methods:** We performed the whole genome sequencing using SARS-CoV-2 positive samples (n=325) collected from February 2020 to the end of June 2021. The number of analyzed samples accounted for 15.4% of the total 2,109 samples identified in our community. Viral lineage was defined by the Phylogenetic Assignment of Named Global Outbreak (PANGO) lineages.

**Results:** We identified 13 types of viral lineages including R.1, P.1, B.1.1.7 (Alpha) and B.1.617.2 (Delta) These virus lineages had distinct periods of expansion and decline. After the emerging of the R.1 lineage harboring E484K variant (designated VOI in Japan), the prevalent B.1.1.214 lineage were no longer identified. The R.1 lineages were temporarily prevalent afterwards, but the influx of B.1.1.7 lineage (designated VOC) led to a decline in R.1. Currently, B.1.1.7 has become dominant after mid-April, 2021.

**Conclusion:** We clearly elucidated the transition and replacement of viral lineage by the community-based analysis. The virus completely replaced by more infectious lineages, therefore, it will be necessary to continue to monitor the VOC and VOI.

## Introduction

The severe acute respiratory syndrome coronavirus 2 (SARS-CoV-2) is spreading worldwide and threatening human health. In countries where vaccination is widely available, the number of infected cases and deaths turned to be decreasing, giving a hope for the virus under the control. Meantime, various types of viral lineage have emerged during the virus evolution. In particular, mutations in receptor binding domain (RBD) of spike protein are of interest for possible changes in the nature of the virus. The several viral lineages have been designated as variant of concern (VOC) or variant of interest (VOI).

On May 31, 2021, World Health Organization (WHO) proposed a new designation, Alpha (B.1.1.7), Beta (B.1.351), Gamma (P.1) and Delta (B.1.617.2) as the four VOCs and Epsilon (B.1.427/B.1.429), Zeta (P.2), Eta (B.1.525), Theta (B.1.525) Lota (B.1.526), Kappa (B.1.617.1) and Lambda (C.37) as the seven VOIs [1]. The U.S. Centers for Disease Control and Prevention (CDC) [2], the European Centre for Disease Prevention and Control (ECDC) [3], the Public Health England (PHE) [4] and National Institute of Infectious Diseases (NIID) in Japan also have their own designations for VOCs and VOIs [5]. These lineages have hallmark mutations in spike protein, which was suggested to affect the transmissibility, immunity, and disease severity [6]. To prevent the spread of SARS-CoV-2, these VOCs and VOIs continue to be under surveillance in many countries.

In January 2021, we started to conduct genomic surveillance of SARS-CoV-2 [7-9]. We previously reported on the SARS-CoV-2 R.1 lineage harboring spike W152L, E484K and G769V mutations [8]. In Japan, the R.1 lineage was first registered in Global Initiative on Sharing All Influenza Data (GISAID) database in November 2020 and showed an increase around January 2021 [8, 10, 11]. Although the NIID in Japan has designated R.1 lineage as a VOC, it is not clear there are associations with increased infectivity and transmissibility [5].

The B.1.1.7 lineage was first identified in the United Kingdom at September 2020 and detected at airport quarantine in Japan at December 2020. The B.1.1.7 was reported to be highly transmissible and increase the disease severity [12, 13]. Actually, the B.1.1.7 spreads rapidly and is identified in 150 countries as of June 30, 2021 [14, 15]. Although B.1.1.7 is reported to be highly transmissible, the transition of other virus lineage in Japan has not been fully elucidated after the influx of B.1.1.7.

In this study, we conducted whole genome analysis of SARS-CoV-2 in 325 samples collected from February 2020 to June 2021 in Kofu, Japan. The lineages other than VOC and VOI were observed until mid-January 2021, afterward, the R.1 lineage was dominant. However, the subsequent influx of B.1.1.7 increased rapidly, suggesting a rapid replacement of R.1 with B.1.1.7. We elucidated that the two major lineages have different infectivity based on genomic surveillance.

## Materials and Methods

### Ethics statement

The Institutional Review Board of the Clinical Research and Genome Research Committee at Yamanashi Central Hospital approved this study and the use of an opt-out consent method (Approval No. C2019-30). The requirement for written informed consent was waived owing to it being an observational study and the urgent need to collect COVID-19 data.

### Nucleic acid extraction

Nasopharyngeal swab samples were collected by using cotton swabs and placed in 3 ml of viral transport media (VTM) purchased from Copan Diagnostics (Murrieta, CA, United States). We used 200 µl of VTM for nucleic acid extraction, performed within 2 h of sample collection. Total nucleic acid was isolated using the MagMAX Viral/Pathogen Nucleic Acid Isolation Kit (Thermo Fisher Scientific; Waltham, MA, United States) as previously described [16].

### Screening of spike protein mutation with TaqMan assay

To detect hallmark mutations found in VOC and VOI, we performed allelic discrimination analysis with a TaqMan assay. We also used a TaqMan SARS-CoV-2 Mutation Panel for detecting spike 69-70 deletion, N501Y and E484K (Thermo Fisher Scientific). We also designed a Custom TaqMan assay (Thermo Fisher Scientific) for detecting SARS-CoV-2 spike protein with the W152L and G769V mutations as previously described [8]. TaqPath 1-Step RT-qPCR Master Mix CG was used as the master mix. The TaqMan Minor Groove Binder probes for the wild-type and variant alleles were labelled with VIC dye and FAM dye fluorescence, respectively.

### Whole-genome sequencing

We studied a total of 335 patients who were infected with SARS-CoV-2 determined by real-time quantitative PCR and/or quantitative antigen test from February 15, 2020 to June 30, 2021 [16-19]. We subjected all these samples to whole genome analysis and successfully obtained 325 sequence data, except for 10 samples with very low viral load.

SARS-CoV-2 genomic RNA was reverse transcribed into cDNA and amplified by using the Ion AmpliSeq SARS-CoV-2 Research Panel (Thermo Fisher Scientific) on the Ion Torrent Genexus System in accordance with the manufacturer’s instructions [8, 9]. Sequencing reads were processed, and their quality was assessed by using Genexus Software with SARS-CoV-2 plugins. The sequencing reads were mapped and aligned by using the torrent mapping alignment program. After initial mapping, a variant call was performed by using the Torrent Variant Caller. The COVID19AnnotateSnpEff plugin was used for the annotation of variants. Assembly was performed with the Iterative Refinement Meta-Assembler [20].

### Clade and lineage classification

The viral clade and lineage classifications were conducted by using Nextstrain [14], and Phylogenetic Assignment of Named Global Outbreak (PANGO) Lineages [21]. The sequences data was deposited in the Global Initiative on Sharing Avian Influenza Data (GISAID) EpiCoV database [22].

## Results

From February 2020 to the end of June 2021, we collected 335 SARS-CoV-2 positive samples determined by RT-qPCR and/or quantitative antigen tests [16-19]. We subjected these samples to the whole genome analysis and TaqMan mutation screening assay. As a result, sequencing analysis could successfully determine viral sequence from 325 individuals, excluding 10 individuals due to the very low viral load. As of June 30, this represented 15.4% of the 2,109 infected individuals identified in our district.

To characterize the viral lineage, the yielded sequence data were analyzed by PANGO lineage [15, 21]. The result showed the 325 samples were classified into 13 types of lineage (B, B.1, B.1.1, B.1.149, B.1.1.284, B.1.1.214, B.1.411, B.1.346, R.1, P.1, B.1.1.220 B.1.1.7 and B.1.617.2) (Table 1).

**Table 1.** Identified virus lineages in the community.

| **Lineage** | **Designation by WHO** | **Designation by CDC (USA)** | **Designation by ECDC (Europe)** | **Designation by NIID (Japan)** | **Number of samples** | **Identified period** |
| --- | --- | --- | --- | --- | --- | --- |
| B | - | - | - | - | 1 | February 2020 |
| B.1 | - | - | - | - | 2 | March 2020 |
| B.1.1 | - | - | - | - | 8 | March to November 2020 |
| B.1.149 | - | - | - | - | 1 | March 2020 |
| B.1.1.284 | - | - | - | - | 20 | July 2020 to January 2021 |
| B.1.1.214 | - | - | - | - | 96 | August 2020 to February 2021 |
| B.1.411 | - | - | - | - | 4 | December 2020 |
| B.1.346 | - | - | - | - | 3 | January 2021 |
| R.1 | - | - | - | VOI | 20 | January to May 2021 |
| P.1 | VOC | VOC | VOC | VOC | 1 | February 2021 |
| B.1.1.220* | - | - | - | - | 3 | April 2021 |
| B.1.1.7 | VOC | VOC | VOC | VOC | 165 | April to June 2021 |
| B.1.617.2 | VOC | VOC | VOC | VOC | 1 | June 2021 |
| Total |  |  |  |  | 325 |  |

VOI, variant of interest; VOC, variant of concern
\* harboring E484K mutation

During the first wave (March to May 2020) in Japan, the B.1 and B.1.1 lineages with spike D614G mutation were predominant [23]. At the second (from July to September 2020) and third waves (from October 2020 to February 2021), the B.1.1.214 (a total of 96 samples) and B.1.1.284 (20 samples) lineages were prevailed. Almost of the B.1.1.214 and B.1.1.284 lineages were identified only in Japan and did not spread to other countries [24].

Afterward, several types of VOC or VOI were identified in our district (Figure 1 and Table 1). The R.1 was first detected in January 2021 and a total of 20 were identified so far (Table 1). Although P.1 was detected in a patient at February 2021 (Figure 1) [9], there was no further spread of P.1 thereafter, suggesting early containment was successful. In the early part of the fourth wave (March to mid-April, 2021), the R.1 and B.1.1.7 lineages coexisted, but after the late part of the fourth wave (after mid-April, 2021), B.1.1.7 became dominant (Figure 1). The first case of B.1.617.2 was found in our community in June, but no further spread of the infection has occurred at this point. The results show that the once potentially dominant R.1 lineage declined rapidly along with the influx of B.1.1.7 lineage.

**Figure 1.**
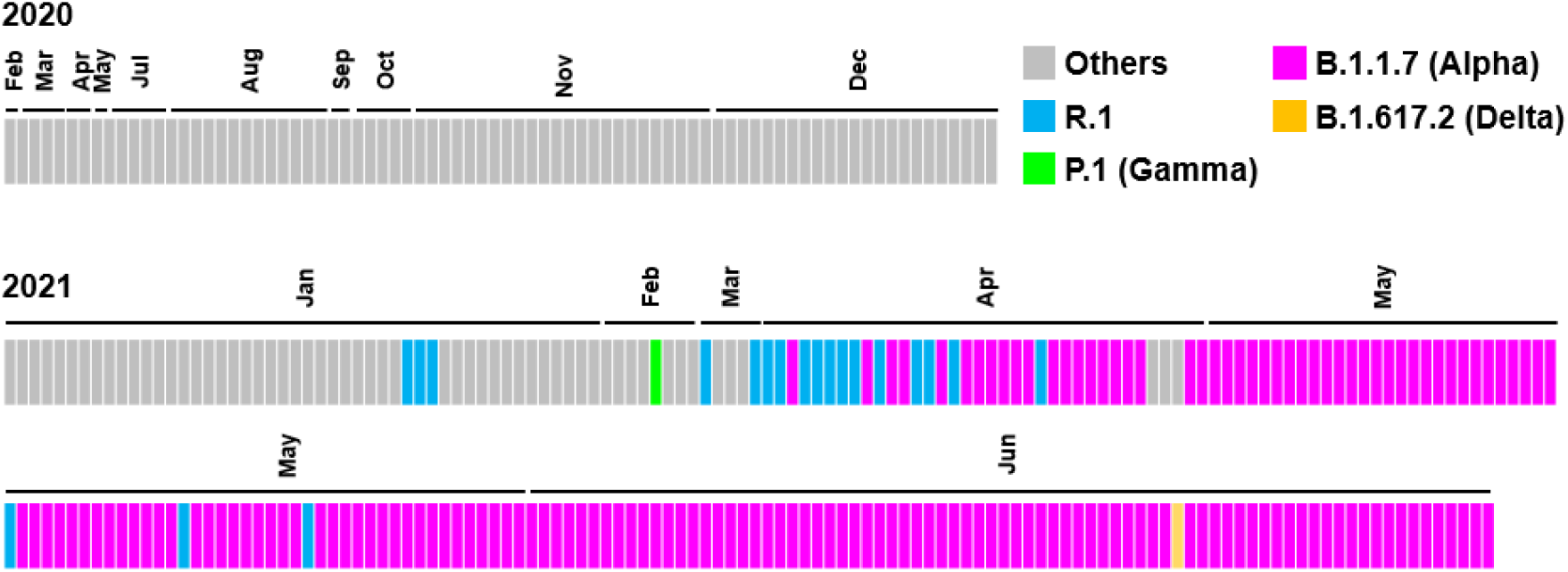
Transition of virus lineages. The data showed the virus lineages identified from February 2020 to the end of June 2020 in a community in Japan. The lineages were analyzed by PANGO lineage. Each box represents a detected lineage and is indicated by R.1 (light blue), P.1 (light green), B.1.1.7 (pink), B.1.617.2 (orange) and others (gray).

## Discussion

The B.1.1.7 has N501Y mutation in RBD of spike protein, binds to the angiotensin-converting enzyme 2 with high affinity and acquires a high transmission rate [25]. This study revealed that the B.1.1.7 rather than the R.1 expanded in Japan during the April to June, 2021. These two lineages were mixed temporally from March to mid-April, 2021, but eventually B.1.1.7 became dominant. Our epidemiological analysis clearly showed the R.1 and B.1.1.7 competed in a community with human migration and that the higher propagating B.1.1.7 lineage survived.

SARS-CoV-2 R.1 lineage emerged rapidly in Japan in January and February 2021 (Figure 1). The R.1 carries the W152L N-terminal domain and E484K in receptor binding domain [8], which have been shown to be of concern for immune escape [26], while there are no reports on its transmission potential. Of note, the fact that the B.1.1.214 lineage was replaced by the R.1 lineage suggests that R.1 possibly has higher transmissibility. This situation is observed by the data of genomic epidemiological study throughout Japanese dataset and in Tokyo [10, 11, 27], indicating current study shows that the situation is not limited to one district.

Recent study showed the relative instantaneous reproduction numbers of the R.1, B.1.1.7, and B.1.617.2 compared to other strains were estimated at 1.256 (range: 1.198-1.335), 1.449 (range: 1.342-1.596), and 1.776 (range: 1.557-2.00), respectively [28]. This data is consistent with the situation observed in our community, suggesting that a virus lineage will be replaced by another lineage with high transmissibility. As of June 30, B.1.1.7 was the predominant in our community, while the first B.1.617.2 strain was identified on June 21. The B.1.617.2 is completely replacing the B.1.1.7 that was rampant and spreading in the United Kingdom [14]. Ito *et al*. estimated that B.1.617.2 lineage will replace others including the B.1.1.7 around mid-July, 2021 in Japan [28]. The COVID-19 mRNA vaccines were shown to be effective against these circulating VOCs [29, 30], however, it is important to continue the genomic surveillance and monitor trends for signs of an increase in the number of infected individuals [31].

In summary, we present a community-based stationary observation of the viral linage replacement. Genomic epidemiological investigations revealed that the whole picture of shifting of VOI and VOC in the community. Prospective whole genome analysis could reveal the linage transitions in real time and the key viral lineage in the infection explosion. Real-time monitoring can provide suggestive insights for public health containment of the virus.

## Data Availability

All data generated or analysed during this study are included in this published article

## Acknowledgments

We also thank Masato Kondo, Ryota Tanaka, Kazuo Sakai, Manami Nagano, Takuhito Fukami, and Ryo Kitamura (Thermo Fisher Scientific) for technical help, all of the medical and ancillary hospital staff for their support, and the patients for their participation. We thank all researchers who share genome data on GISAID (http://www.gisaid.org).

